# Consequences of Mismatch, Misalignment and Rotation of Toric Intraocular Lenses in Refractive Cataract Surgery Part 2. Avoiding Flip Flops

**DOI:** 10.1101/2020.09.30.20203380

**Authors:** Samir I Sayegh

## Abstract

**Purpose:** To show current approaches for overcorrecting astigmatism and “flipping” its axis need be reconsidered in light of methods we introduce that take into account both mismatch and misalignment of the toric intraocular lens (tIOL) with respect to the astigmatism to be corrected at the time of cataract surgery.

**Setting:** Private Practice and Research Center. The EYE Center. Champaign, IL, USA.

**Design:** Formal Analytical Study

**Methods:** In the most common surgical situation where both mismatch and misalignment exist, we present an analysis of the point at which overcorrection and undercorrection residuals coincide, yielding a simple but powerful methodology to predict the optimal degree of overcorrection with a tIOL. The method is illustrated for tIOLs used in surgical practice.

**Results:** The minimum astigmatism appropriate to overcorrect with a tIOL is given by, 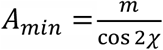,where *m* is the midpoint threshold used by “split-the-difference” algorithms and *χ* is the estimate of tIOL misalignment due to all causes. Correspondingly, the maximum overcorrection, Ω_*max*_, that should be attempted is 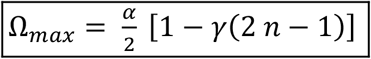 where 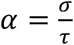 is the *dioptric step at the corneal plane*, with σ = *H* − *B*, where *H* = *n* σ is the cylinder of the overcorrecting tIOL and *B* = (*n* − 1)σ is the cylinder of the undercorrecting tIOL, both at the IOL plane, *τ*is the toricity ratio and *γ* relates to the angle of misalignment *χ* by 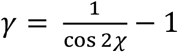 which can be approximated by 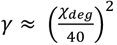. Ω _*max*_factors elegantly in the product of 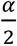, the (ideal) midpoint correction for perfect alignment, by the bracketed term, representing the percent reduction of the ideal value in a realistic surgical situation with estimated misalignment *χ*. To illustrate: an eye of average dimensions 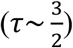 and tIOLs from major manufacturers 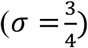, with *A* = 2.35 D dictating n = 5. For a misalignment of 10°Ω_*max*_≅ 0.10 *D* is the maximum overcorrection that should be accepted, significantly smaller than the midpoint 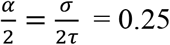 D, recommended by many current tIOL calculators.

**Conclusion:** An optimal method is presented for the selection of an overcorrecting tIOL at the time of refractive cataract surgery with improvement over current tIOL calculators’ methods.

## INTRODUCTION

Toric calculators have historically avoided overcorrection sometimes based on misunderstandings of the optical principles underlying image formation and perception. More recently, a majority of toric calculators have migrated to a “split-the-difference” strategy where overcorrection is accepted if the residual provided by the overcorrecting tIOL is less than that of the undercorrecting one^12^. However, the decision of overcorrecting at an apparently favorable residual and thus “flipping” the axis may be more subtle than originally considered.

In part one of this trilogy, we established that the *angle of doom*, Δ, where astigmatism correction is nullified by misalignment can vary depending on the degree of mismatch between the cylinder value of the correcting tIOL and that of the astigmatism it is intended to correct. We gave a rigorous treatment and precise results including very solid estimates that can be computed “on the fly” in a clinical surgical setting. Specifically we showed that **Δ ≈ 30 − 15 *ω***with ***ω***being the relative overcorrection with respect to the astigmatism to be corrected and similarly **Δ ≈ 30 + 15 *μ*** with ***μ*** being the relative undercorrection^3^. The implication is that residual astigmatism increases faster in the presence of overcorrection as compared to undercorrection. At zero misalignment with the astigmatism it is intended to correct, an overcorrecting tIOL residual, initially smaller than a corresponding undercorrecting one, will rise faster with rising misalignment. As misalignment increases, it will reach then exceed the slower rising undercorrecting residual, at which point it will cease to be the optimal choice. Because misalignment due to a variety of reasons is inevitable, the best strategy for potential overcorrection is to select the overcorrecting astigmatism, not at zero alignment, which almost never occurs, but rather at the largest degree of misalignment where it is still favorable as compared to the corresponding undercorrection.

We address this problem analytically with the same methodology used in part one and give a full solution with exact results as well as useful rules of thumb that can be considered for clinical practice and surgical planning. We then provide examples of several current toric calculators where the split-the-difference approach is consistently used, possibly resulting in less than optimal choices of a tIOL and provide the corresponding optimal solutions.

## METHODS

### Astigmatism Representation

We use three types of simple diagrammatic representations to guide the reasoning and the derivations that follow. A triangle representation (Figure [1]) in which the residual R is the length of the third side of a triangle with sides A and L, where A represents the astigmatism to be corrected at the corneal plane, and L is the tIOL cylinder, also at the corneal plane. There is a variable misalignment, λ, between L and A which is represented by 2 λ in the triangle representation, as is customary. The second representation is an “astigmatism levels” representation where each astigmatism is represented by a horizontal line as shown in Figure [2]. Finally, we plot (Fig [3]) the values of the residual as a function of the angle of misalignment λ for undercorrection and overcorrection and consider the angle λ = *χ* where these plots intersect.

Applying the law of cosines to the ALR triangle of Figure 2

**Figure 1.**
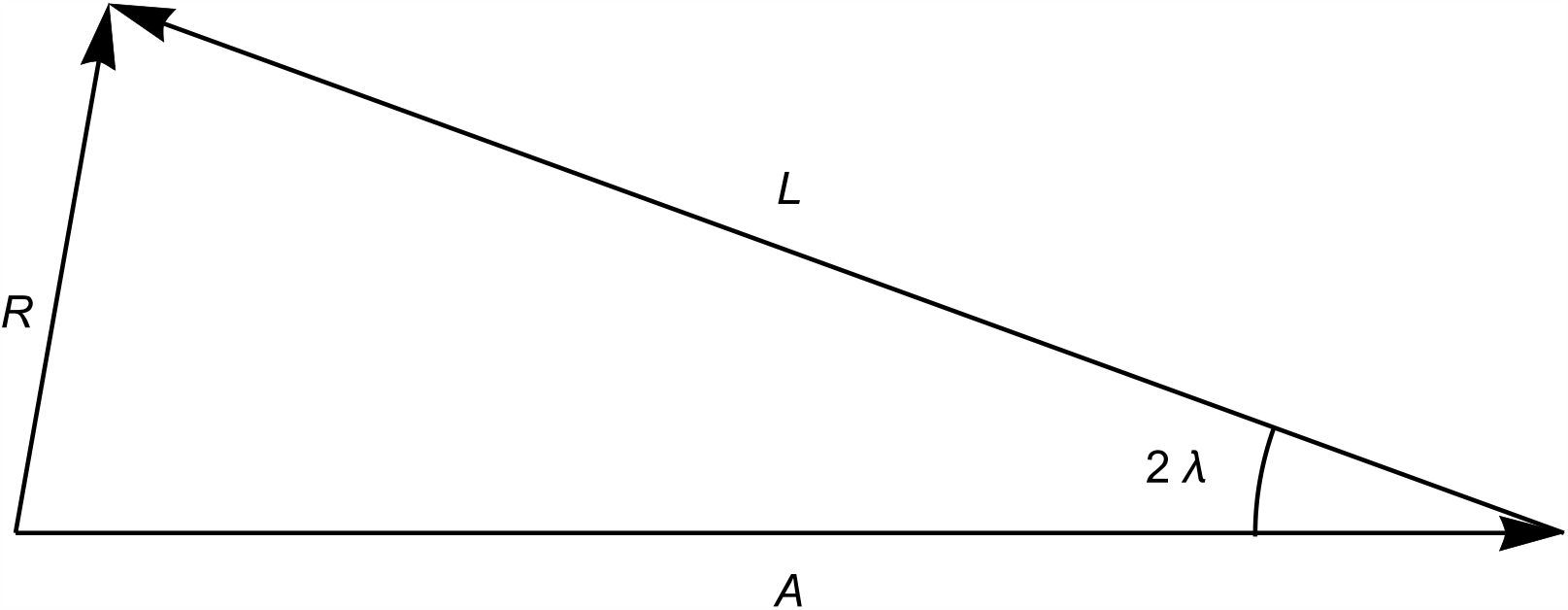
A triangle representation of astigmatism A with an attempted correction L that has rotated by an angle λ with respect to the intended meridian, with a residual astigmatism R. The geometric representation and residual calculation are based on a triangle with an angle 2λ between the sides, A and L. From elementary geometry an “SAS” (side angle side) triangle is uniquely determined and therefore both the residual astigmatism and its meridian are easily computable.

**Figure 2.**
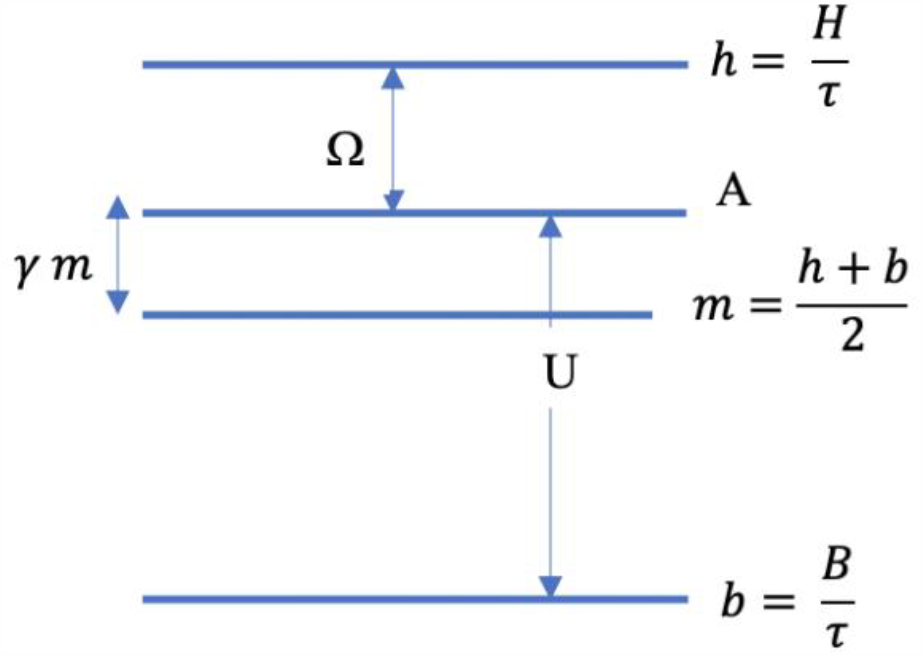
Diagrammatic astigmatism “levels” representation at the corneal plane. A is the astigmatism present and to be corrected at time of surgery, H and h are the overcorrecting (“high”) tIOL cylinder values at the tIOL and corneal planes respectively, related by *τ*, the toricity ratio, as indicated. Similarly, B and b are the undercorrecting (“below”) tIOL cylinder values at the tIOL and corneal planes, respectively. m is the midpoint, or average of h and b. Ωis the amount of overcorrection when H (and correspondingly h) is chosen and U the amount of undercorrection when (B and correspondingly b) is chosen. γis the fractional increment of A with respect to m.

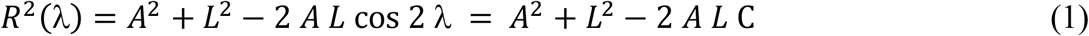

where we set cos 2λ = *C*.

Figure 2 illustrates that the astigmatism, A, to be corrected at the corneal plane is bracketed or “sandwiched” between a *high* value, *h* diopters, and a “be*low*” value, *b* diopters, both at the corneal plane, corresponding to overcorrecting tIOLs with tIOL plane values *H* diopters and *B* diopters, respectively. IOL plane values are related to corneal plane values by the toricity ratio, *τ*. *H* and *B* are assumed to be separated by a discrete value or “step” of *σ* diopters (we will refer to this discrete finite separation of *H* and *B* as the “first quantization of astigmatism correction”). Finally, we designate the ratio 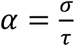 as the *dioptric step at the corneal plane*. In other words, we have,

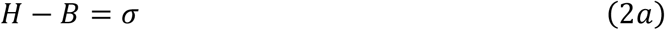

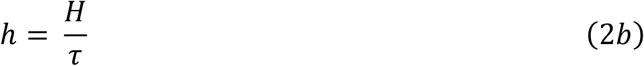

and,

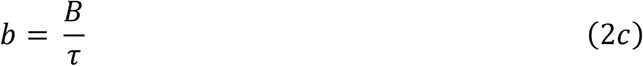

Combining 2a, 2b and 2c we obtain,

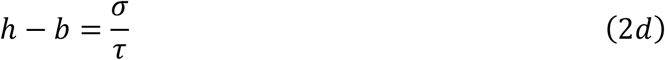

We also introduce the midpoint, *m*,

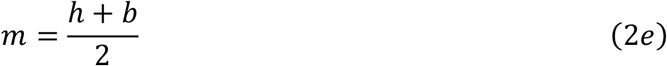

and the value *n*,

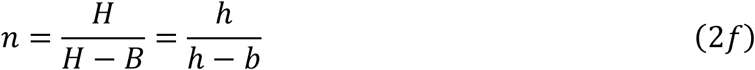

which is not necessarily, but often turns out to be, an integer.

Combining 2a and 2f, we can write,

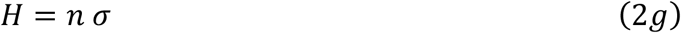

And similarly,

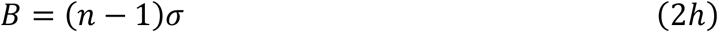

And combining with 2a and 2b and 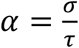 we can write,

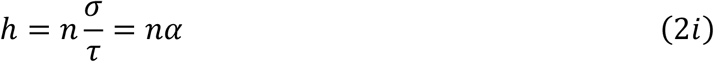

And,

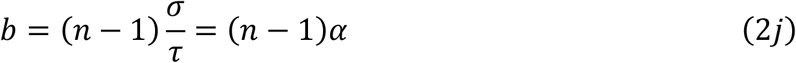

From which we have,

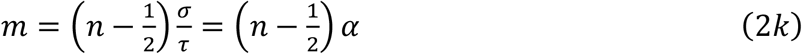

Note also that,

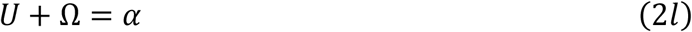

If we express the various astigmatic quantities in units of 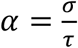 we can simply visualize the levels of overcorrection, undecorrection tIOLs, and midpoint as expressed by *n, n* − 1, and 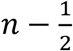

## RESULTS

### When does it make sense to overcorrect astigmatism and “flip” the axis?

Using Equation 1 we can calculate the residual astigmatism for any under or overcorrection of an astigmatism, A, and display their variation as a function of the degree of misalignment, λ.

Figure [3] represents an example of the rising degree of residual astigmatism as a function of the angle of misalignment for both an overcorrection and an undercorrection with the intersection point at a *crossing angle χ* occurring between 4 and 5 degrees of misalignment. While initially smaller, the residual overcorrection ceases to be advantageous beyond *χ*.

We can now formalize this approach to arrive at exact results as well as useful clinical approximations.

The residual astigmatism resulting from undercorrecting A diopters of astigmatism with *b* diopters, (*b* < *A*), will satisfy equation (1) and thus we will have:

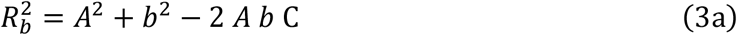

Similarly, the residual astigmatism resulting from an overcorrection of A diopters with *h* diopters, (*h* > *A*), will also satisfy equation (1) and thus we will also have:

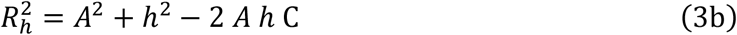

Equal residuals for over and undercorrection occur at a *crossing angle* λ = *χ* and is obtained by equating (3a) and (3b). Resulting in

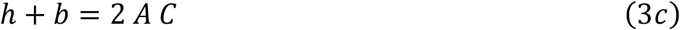

Writing in terms of 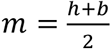, the midpoint between *h* and *b*, we now have for the minimal astigmatism to be overcorrected,

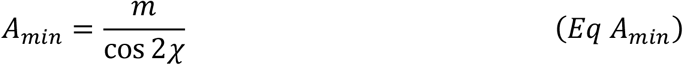

Which can also be written as,

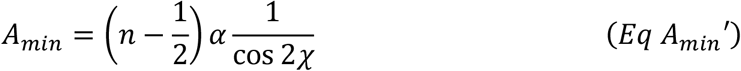

Since 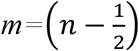α.

Note that *A*_*min*_≥ *m* where the midpoint 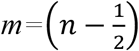α is the typical value used for “split the difference” algorithm. Since cos 2*χ* ≤1, *A* will be larger than *m* and will continue to grow with increasing *χ* throughout the clinically realistic range. To illustrate, for astigmatism 1.30 D, bracketed by *b* = 1.00 D and *h* = 1.50 D, a split-the-difference approach would suggest that any *A* above = 1.25 D should be overcorrected. However, if we take *χ* = 10°for example we then have, 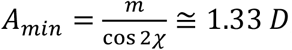, and astigmatism up to 1.33 D should continue to be undercorrected.

To express the crossing angle as a function of A, *σ*, and *τ*we can invert equation *Eq A*_*min*_,

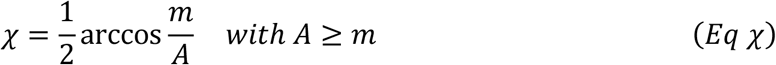

For a given eye characterized by (*A, τ*) and a given manufacturer’s tIOLs offerings characterized by (*n, σ*) the crossing angle is determined by (*Eq χ*).

Using equation (2a) to (2l) we can also express *χ* in terms of *H*, Ω, *B, U*

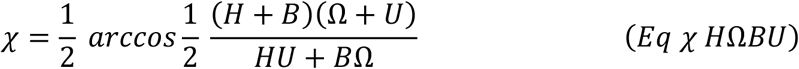

There are two clinically important questions that are answered by (*Eq χ*) and (*Eq A*_*min*_): (*Eq χ*) answers the question of determining the crossing angle of over and undercorrecting tIOLs selected from a sequence of tIOLs characterized by (*n, σ*) for astigmatism *A* in an eye characterized by (*A, τ*). (*Eq χ H*Ω*BU*) allows for this calculation to be done without direct knowledge of A but rather through values returned by existing calculators. Once that angle is determined it can be decided if such an angle is adequate or too small in order to accept or reject the corresponding degree of overcorrection. For example, in Figure 3 an astigmatism of 3.90 D is to be corrected and, at the corneal plane, the overcorrecting and undercorrecting tIOL powers are given by 4.11 D and 3.60 D respectively, yielding 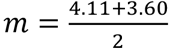 and 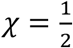arccos 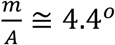consistent with the graphical representation in Figure 3. The clinical question then is whether the preoperative and intraoperative measurements including all estimates of astigmatism (anterior, posterior, SIA, cyclotorsion, alignment, other) and potential postoperative rotation combine to about 4 degrees or less, in which case the overcorrection should be accepted and otherwise the undercorrection should be preferred.

**Figure 3.**
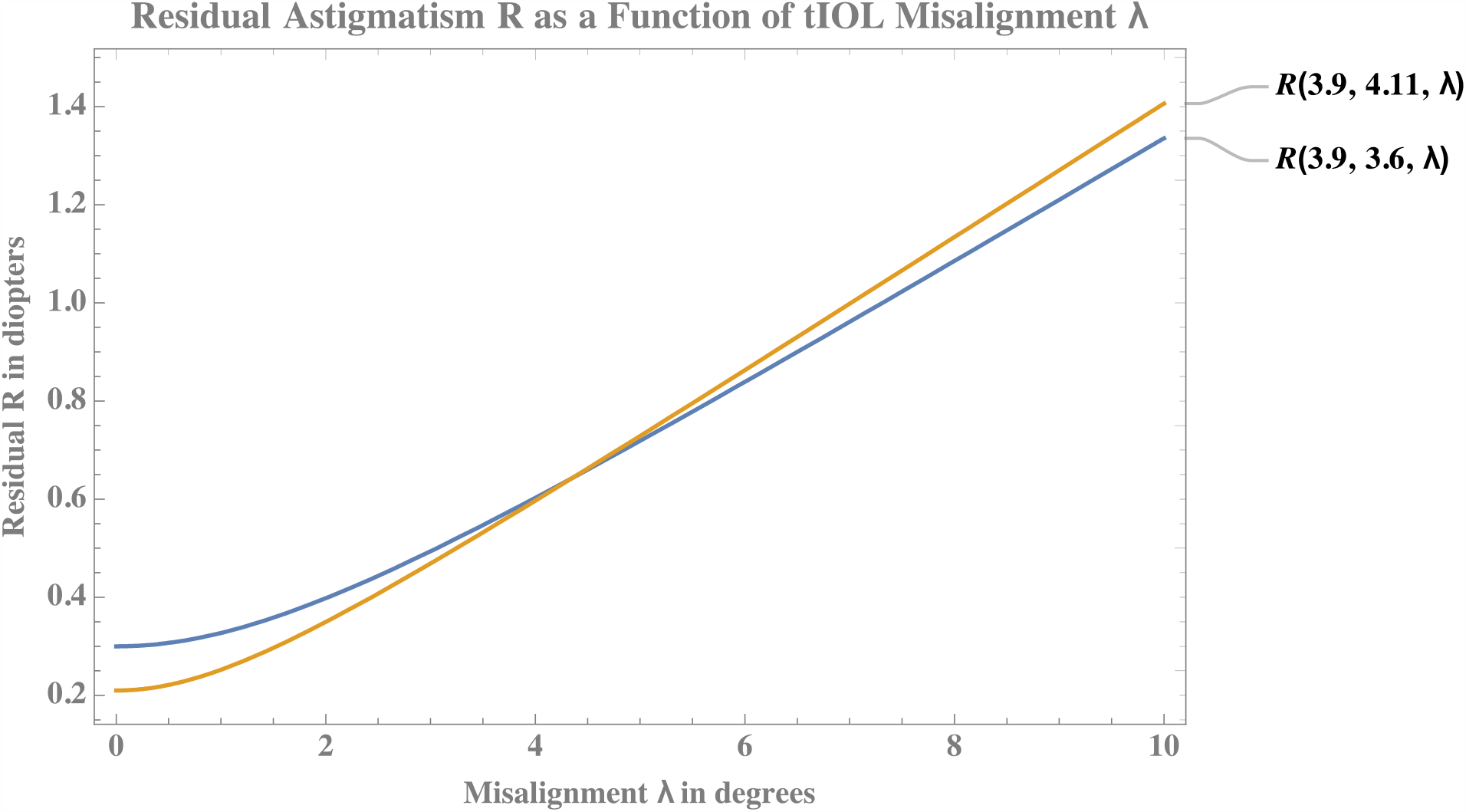
Astigmatism A=3.9 D with an initially favorable overcorrecting tIOL of 4.11D (residual 0.21D) and an undercorrecting one of 3.60 D (residual 0.30 D), all at the corneal plane. Beyond misalignment by an angle λ= χ∼ 4 degrees, the residual associated with overcorrection equals then starts exceeding the residual of the undercorrection. R(A,L,γ) represents the residual for astigmatism A corrected by L, with misalignment γ. See Equation (1).

(*Eq A*_*min*_) on the other hand, requires an input of an estimated crossing angle *χ* in addition to the other parameters. That value of *χ* is presumed to be the estimate of the degree of misalignment from all causes. A value of *A*_*min*_ is then computed at or below which only undercorrection should be accepted. As an example, take the (very optimistic!) value of *χ* = 5°and using the same value of *m*, we obtain 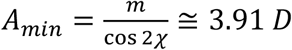. Signifying that in the presence of a misalignment of 5°, the least astigmatism to be corrected should be 3.91 D, a value consistent with the previous example and illustrated in Figure 3. It is possible to graph corresponding values of A and *χ* and use such graph as a quick look up table. See Appendix B.

Sometimes it is more convenient to deal with the degree of overcorrection directly, since for some calculators the value of total corneal astigmatism is not explicitly presented (for example because an undisclosed contribution of posterior astigmatism is included).

As indicated in Figure 2, the overcorrection Ω is given by Ω = *h* − *A* and we can easily derive, using *Eq A*_*min*_:

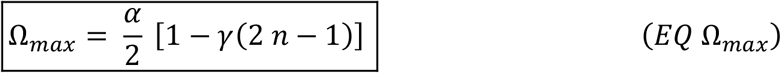

*with*,

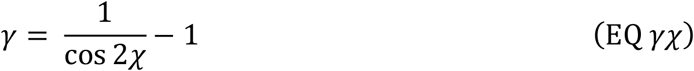

In Appendix *A* we show that *γ* has a simple interpretation as well as the useful approximation:

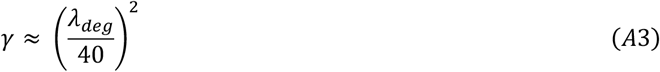

Note that 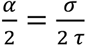 being the midpoint correction between *h* and *b*, Ω _*max*_ *is lesser than that midpoint value for any positive values of A and χ*. The reduction from that midpoint threshold is proportional to A (via *n*) and to *χ*^2^ for most realistic values of the misalignment *χ*. In other words, *the larger the misalignment and the higher the amount of astigmatism to be corrected, the smaller the allowed overcorrection*, the optimal degree of overcorrection that should be sought potentially reaching zero or a negative value, precluding any consideration of overcorrection.

One can also see that in the limit *A* → 0 or *χ* → 0, we have 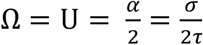, the midpoint or “split-the-difference” solution.

As an example consistent with the previous ones we choose once again the value of *χ* = 5°and use the approximation from equation *A*3 to compute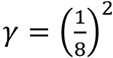 and with *n* = 8, *σ* = 0.*7*5 D and *τ*≅ 1.46 we have 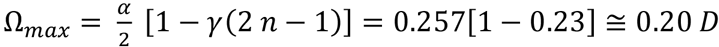. This answer is consistent with the one obtained following the *A*_*min*_method since we easily verify *A*_*min*_+ Ω_*max*_ = *h*. (3.91+0.20 = 4.11). This signifies that no larger overcorrection than 0.20 D should be accepted in this context if the misalignment is estimated at *χ* = 5°. We can observe that the bracketed value represents the percent reduction of the admissible overcorrection as compared to the “split-the-difference” 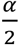 value corresponding to the perfect alignment case *χ* = 0°.

We now turn to a systematic presentation of the method to be followed to determine optimal overcorrection.

### Methods to determine optimal degree of overcorrection

One can use either equation (*Eq A*_*min*_) or (*EQ* Ω_*max*_) to determine the optimal degree of overcorrection for a given estimate of misalignment *χ*, at which the residual curves intersect and overcorrection is no longer advantageous. The preferred method will depend on personal intuition and on whether the surgeon is performing her or his own calculations “from scratch” or relying on a specific toric calculator that may or may not provide easy access to the parameters needed.

We illustrate the method using (*Eq A*_*min*_) and (*EQ* Ω_*max*_) This gives rise to a method or algorithm to optimize the choice of tIOL by determining the maximum allowable overcorrection. The steps outlined can be done *de novo* or read or deduced from values provided by an existing toric calculator, even though that calculator may be recommending a suboptimal choice.

### *de novo* Methods

The input needed is

1. biometric specification for an eye and planned incisions with corresponding SIA allowing for total corneal astigmatism *A* and toricity ratio ***τ*** to be determined. This information depends on measurements performed on the individual EYE.
2. a sequence or subsequence (minimum 2) of tIOLs that allow for the step *σ*, to be specified in diopters, as well as *n, n being a single value or a sequence of possible values*. This is done via equations (2a) and (2f). This information depends on tIOLs made available by a MANUFACTURER.
3. an estimate or computation of the crossing angle of misalignment λ = *χ* due to all causes. This estimate depends on reliability of instrumentation and models used to estimate various measurements and previous experience all to be assessed by the SURGEON.

### *de novo A*_*min*_

One can then apply “Method *A*_*min*_” that proceeds as follows:

1. Determine *σ* by selection of manufacturer’s sequence or subsequence of tIOLs (Eq 2a).
2. Determine *τ* by measuring the eye’s axial length, K values, and any other biometric variables and using manufacturers’ recommendations or/and using an appropriate formula or nomogram^45^. Average *τ* is about 1.30 −1.50 depending on type of IOL and increasing with increasing axial length and corneal curvature.
3. Determine 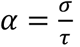 the *dioptric step at the corneal plane*.
4. For astigmatism A to be corrected at the corneal plane, calculate *n* by determining *H* as the smallest value of a tIOL larger than the product *τA*, and determining *B* as the largest value of a tIOL smaller than the product *τA*. Then use 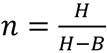. Most manufacturers often provide a sequence given by *H* = *n σ*,where *n* takes integer values, and we then simply have 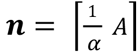. Select the corresponding candidate tIOLs, B = (*n* − 1) *σ* and H = *n σ*, undercorrecting and overcorrecting A. See for example Table 1 for an example of tIOLs sequence.
5. Estimate *χ* based on surgical experience or based on the estimate of all contributions to misalignment in a surgical case, then calculate 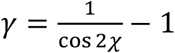 or use the excellent approximation 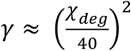
6. Substitute the values obtained in steps 1 through 5 in 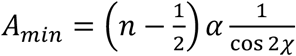
7. Overcorrect A with *H* if and only if *A* > *A*_*min*_, otherwise choose *B* = *H* − *σ*

### *de novo* Ω_*max*_

One can similarly apply “Method Ω_*max*_” as follows:

1. Determine *σ* by selection of manufacturer’s sequence or subsequence of tIOLs.
2. Determine *τ* by measuring the eye’s axial length, K values and any other biometric variables and using manufacturers’ recommendations or/and using an appropriate formula or nomogram^45^. Average *τ* is about 1.30 −1.50 depending on type of IOL and increasing with increasing axial length and corneal curvature.
3. Determine 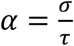 the *dioptric step at the corneal plane*.
4. For astigmatism A to be corrected at the corneal plane, calculate *n* by determining *H* as the smallest value of a tIOL larger than the product *τA*, and determining *B* as the largest value of a tIOL smaller than the product *τA*. Then use 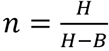 Most manufacturers have a sequence given by *H* = *n σ, n integer* and we then simply have 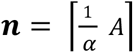. Select the corresponding candidate tIOLs, B =(*n* − 1)*σ* and H= *nσ*, undercorrecting and overcorrecting A. See for example Table 1 for an example of tIOLs from a specific manufacturer.
5. Estimate *χ* based on surgical experience or based on the estimate of all contributions to misalignment in a surgical case, then calculate 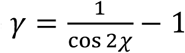 or use the excellent approximation 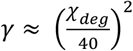
6. Substitute the values obtained in steps 1 through 5 in 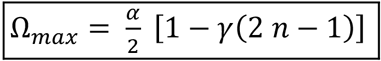
7. Overcorrect A with *H* if and only if 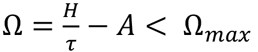, otherwise choose *B* = *H* − *σ*

**Table 1.** The T_n_sequence of tIOLs characterized by a sequence T_n_=0.75 (n− 1) at the tIOL plane and corresponding cylinder values at the corneal plane varying with toricity ratio. Here *σ* =0.75 D and n={3,4,5,6,7,8}

| T | IOL Plane | Avg Cornea Plane | Min Cornea Plane | Max Cornea Plane |
| --- | --- | --- | --- | --- |
| 3 | 1.50 | 1.03 | 0.83 | 1.25 |
| 4 | 2.25 | 1.54 | 1.25 | 1.88 |
| 5 | 3.00 | 2.06 | 1.67 | 2.50 |
| 6 | 3.75 | 2.57 | 2.08 | 3.13 |
| 7 | 4.50 | 3.08 | 2.50 | 3.75 |
| 8 | 5.25 | 3.60 | 2.92 | 4.38 |
| 9 | 6.00 | 4.11 | 3.33 | 5.00 |

Note that the astigmatism, A, to be corrected is the corneal astigmatism resulting from *all contributions*, including anterior astigmatism, posterior astigmatism and astigmatism induced by surgical incisions. It is typically equivalent to the residual astigmatism that would result in case a non-toric intraocular lens is used. A number of modern toric calculators display such a value along options for tIOL choices with their corresponding residuals. The present work assumes an appropriate methodology for computing such a value for A. It also emphasizes that its importance stems in part from the lack of an accepted standard and the variety of methods that yield different results, introducing uncertainties that contribute to both mismatch and misalignment.

A number of calculators will provide a set of values from the ones shown in Figure 2 with the relations as described in 2a to 2l and Appendix C. We are mainly interested in overcorrection Ω, undercorrection *U* and corresponding tIOLs cylinders, H and B, at the IOL plane. We will refer to the (many) calculators that provide these values as *H*Ω*BU calculators* and we show the simple algorithms to use in those cases to determine *A*_*min*_ and Ω_*max*_ as follows.

### Methods for *H*Ω*BU calculators*

*H*, Ω, *B, U* are given in diopters.

#### Based on Ω

1. Determine ***α*** = Ω + *U*, the *dioptric step at the corneal plan*
2. Calculate *n* by 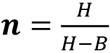
3. Estimate ***χ*** based on surgical experience or all contributions to misalignment, then calculate 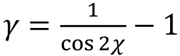 or use 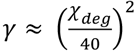
4. Substitute the values obtained in 1) to 3) in 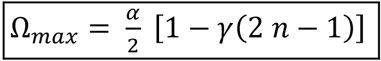
5. Overcorrect with *H* if and only if Ω < Ω_*max*_, otherwise choose *B*

#### Based on A

1) Through 3) as above

4) 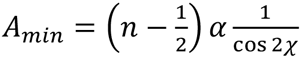

5) Overcorrect *H* if and only if *A*_*min*_ < *A*

(*with A provided* B*y calculator or evaluateD* B*y* 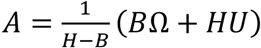, else use *B*

### Common tIOLs and examples

A common situation with tIOLs in increments of α diopter at the IOL plane is that of a “*T*_*n*_” sequence with 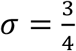 diopter (ZCT, ZCU from JJ and Alcon SN6ATwhere *T*_n_ = *σ* (*n* − 1) are examples encountered later in the paper). A very common clinical scenario is the one where n = 3, resulting in a T3 / T4 pair choice, for which T3 corrects 1.50 diopters and T4 corrects 2.25 diopters respectively at the IOL plane with an approximate correction for an average eye of 1 diopter and 1.50 diopters at the corneal plan, which assumes a toricity ratio of about 1.50. An eye of axial length 24 mm and average K of 44 diopters would correspond to such a toricity ratio but depending on the toricity ratio the corresponding cylinder at the corneal plane may potentially vary by less than half a diopter for a T3 and up to or exceeding 2 diopters for higher *T*_*n*_ as indicated in Table [1].

For this common situation we now have:

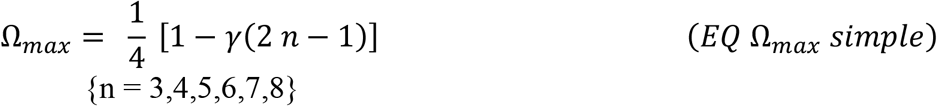

For γ = 0, the trivial *no misalignment* case, the expected value of 1/4 diopter is the “midpoint” value where overcorrection and undercorrection yield equal residual astigmatism. Generally, however, γ will be different than zero and the result will also depend on the degree of attempted astigmatism correction via *n*.

Consider the example of an astigmatism of 1.30 diopter, potentially correctable by a T3 tIOL with cylinder 1.00 at the corneal plane, or a T4 with 1.50 diopter at the corneal plane. The midpoint is 1.25 diopter and 1.30 is in excess of 0.05 diopters of the midpoint. This corresponds to a fraction γ = 0.05/1.25 = 1/25. Substituting in equation(*Eq* λ_*deg*_) we obtain an angle of 8 degrees. This means that overcorrection will continue to be advantageous up to a misalignment/rotation of 8 degrees. If we anticipate a misalignment/rotation of 10 degrees (corresponding to γ = 0.064) it would be preferable, then, to undercorrect. Indeed, proceeding with “Method Ω_*max*_” and substituting γ = 0.064 and n = 3 in equation Ω_*max*_ we obtain Ω_*max*_ = 0.17 diopter. The maximum overcorrection that should be allowed in this case should thus be 0.17, and the 0.30 undercorrection would be preferable to the 0.20 overcorrection.

Lookup tables or nomograms can be easily generated. For example, Table [2] can serve as a practical reference for an “average eye” as indicated by the first row with entries of 0.25 D.

**Table 2.** The first column gives the estimated misalignment/rotation angle of the toric IOL. The subsequent columns give, in diopters and for each pair of undercorrecting-overcorrecting tIOLs,the maximum amount of astigmatism that should be considered for overcorrection for each T_n_pair, for an “average” eye.

|  | T3-T4 | T4-T5 | T5-T6 | T6-T7 | T7-T8 | T8-T9 |
| --- | --- | --- | --- | --- | --- | --- |
| 0 | 0.25 | 0.25 | 0.25 | 0.25 | 0.25 | 0.25 |
| 5 | 0.23 | 0.22 | 0.22 | 0.21 | 0.20 | 0.19 |
| 10 | 0.17 | 0.14 | 0.11 | 0.07 | 0.04 | 0.01 |
| 15 | 0.06 | NA | NA | NA | NA | NA |

The first column gives the estimated misalignment/rotation angle of the toric IOL. As pointed out previously, all sources of misalignment errors need be included (uncertainly on axis measurement, SIA estimate error, posterior astigmatism uncertainty, marking, cyclotorsion, rotation of tIOL, etc…). The first row gives a constant value of 0.25 D indicating a perfect alignment, hardly a realistic assumption in clinical practice. Many surgeons can consider using an estimate around 10 degrees but both larger and smaller values can be considered based on a meticulous assessment of methods and historical surgical results. For 10 degrees, and even for much more optimistic estimates of 5 degrees, it is clear that the now conventional “split the difference” approach is far from ideal, especially as we correct higher degrees of astigmatism. The last row has only one minimal valid entry to T3-T4, and none corresponding to correction of higher degrees of astigmatism. This means that if the estimated misalignment is 15 degrees, only T3-T4 pair may potentially benefit from any overcorrection at all and only if this overcorrection is equal to or lesser than 0.06 D (for example it is still better in this case to undercorrect by 0.40 then to overcorrect by 0.10 D). All other pairs of tIOLs would always favor the undercorrecting tIOL no matter the discrepancy between the residuals. This recommendation is unlike, and preferable to, that of current toric calculators.

For 8 degrees of misalignment and using the approximate expression of *γ* we get 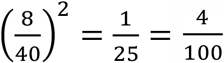 This value of *γ* multiplied by 2 *n* − 1 will be reduced from 1 to yield a percent reduction of the split-the-difference value 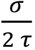. For n = 7 we have a reduction of 52%, in other words the acceptable overcorrection should be less than half the one usually accepted for “split the difference” approaches.

### Further Examples from Various Toric Calculators

We now show a sequence of examples from different manufacturers. The purpose of showing these examples is to demonstrate the methods presented in this paper in a practical useful way and also to demonstrate that the leading manufacturers of tIOLs have broadly adopted a split the difference approach where the overcorrection is always accepted until it becomes equal or superior to the undercorrection. As we discussed, this approach assumes zero misalignment and should be replaced by the methods presented that will incorporate a realistic value of potential misalignment.

#### PhysIOL split-the-difference with/without posterior corneal astigmatism inclusion

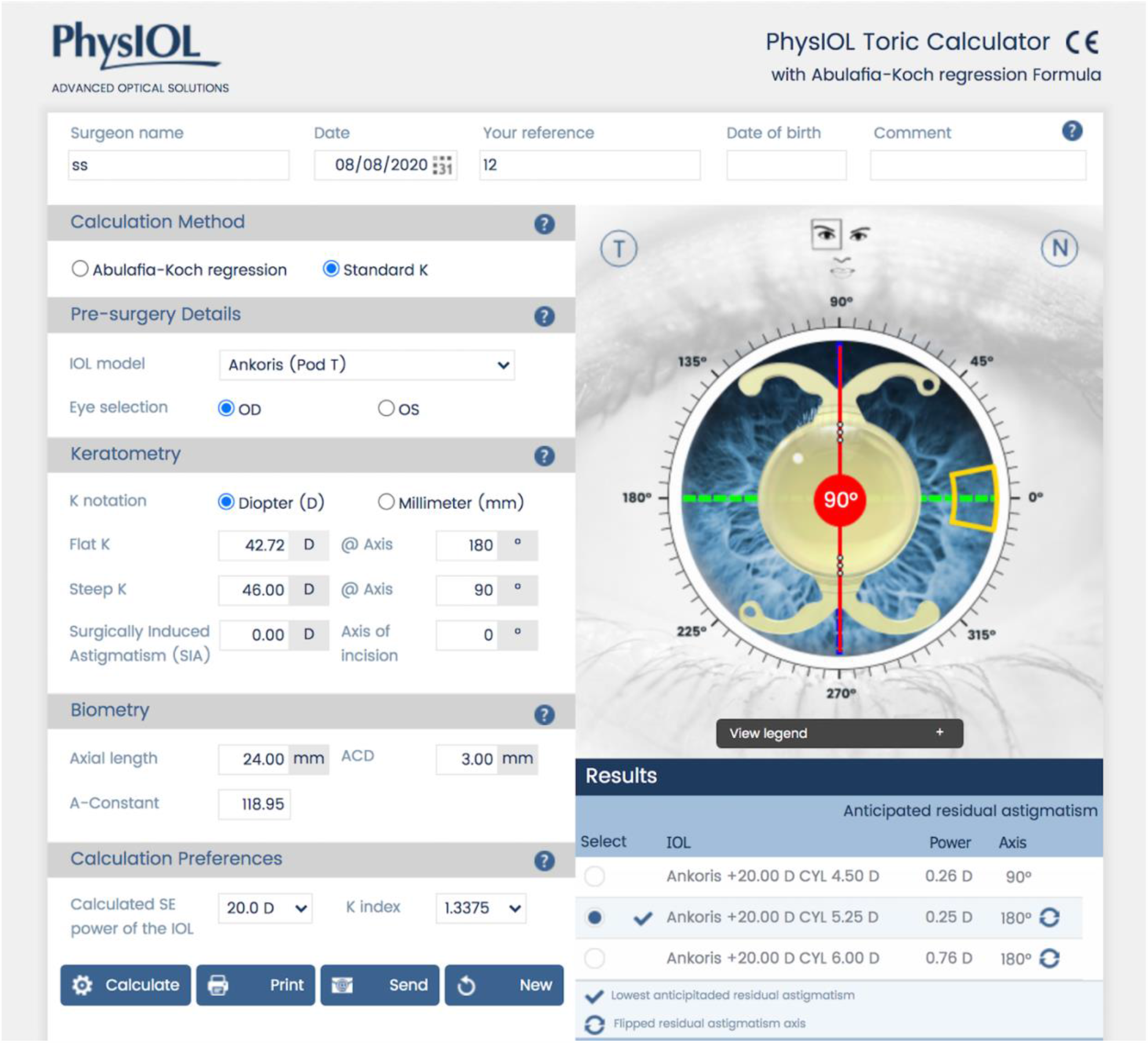

In this standard algorithm (that assumes no posterior astigmatism contribution) we have A = – 42.72 = 3.28 D. This is a ***H*Ω*BU*** calculator and we can apply the corresponding method Here (*H*, Ω, *B, U*) = (5.25,0.25,4.50,0.26) diopters

1. Determine ***α*** = Ω + *U* = 0.25 + 0.26 = 0.51, the *dioptric step at the corneal plan*
2. Calculate *n* by 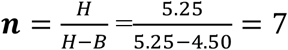
3. Estimate ***χ*** based on surgical experience or all contributions to misalignment, then calculate 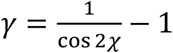 or use 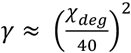. We take ***χ*** = **8**°. 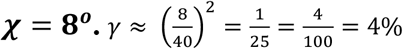
4. Substitute the values obtained in 1) to 3) in 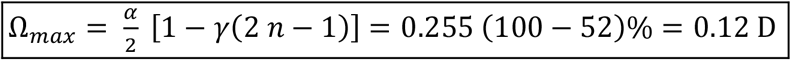
5. Overcorrect with *H* if and only if Ω < Ω_*max*_, otherwise choose *B*. Since Ω = 0.25 *D* and Ω_*max*_ = 0.12 *D* we clearly have Ω > Ω_*max*_ and overcorrection should be avoided and undercorrection selected.

Here we can also apply the *A*_*min*_ method

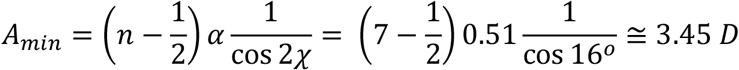

This is larger than the value of 46 – 42.72 =3.28 D of the total astigmatism as entered^1^, and again the overcorrection is not recommended in this circumstance.

Note that we can verify *A*_*min*_ + Ω_*max*_ ≅ *h* = *nα* as we would expect. Indeed, we have *A*_*min*_+ Ω_*max*_≅ 3.45 + 0.12 = *7* × 0.51 = 3.5*7*. The two methods give the same result, as should be expected.

Note that the value suggested by our method, 0.12 D, is well below the value of 0.25 D that has been accepted by the PhysIOL calculator.

We can also determine the crossing angle for the values chosen by the calculator by applying *Eq* (*χ H*Ω*BU*). We find *χ* ≅ 1.6°. If the surgeon is willing to accept the recommendation given, misalignment would need to be no more than about 1 degree. This may be an unrealistic expectation in most surgical cases.

We now look at an example from the same calculator with the Abulafia-Koch correction. We first make the observation that the methods that add a contribution of posterior astigmatism not only modify the estimate of the magnitude of the astigmatism to be corrected but also its meridian. While it is not the intention of this paper to address this topic we point out that the discrepancy between the axis when posterior astigmatism is taken into account and when it is not can introduce an uncertainty of several degrees on the final axis of the astigmatism to be corrected, thus further justifying the need for the methods presented.

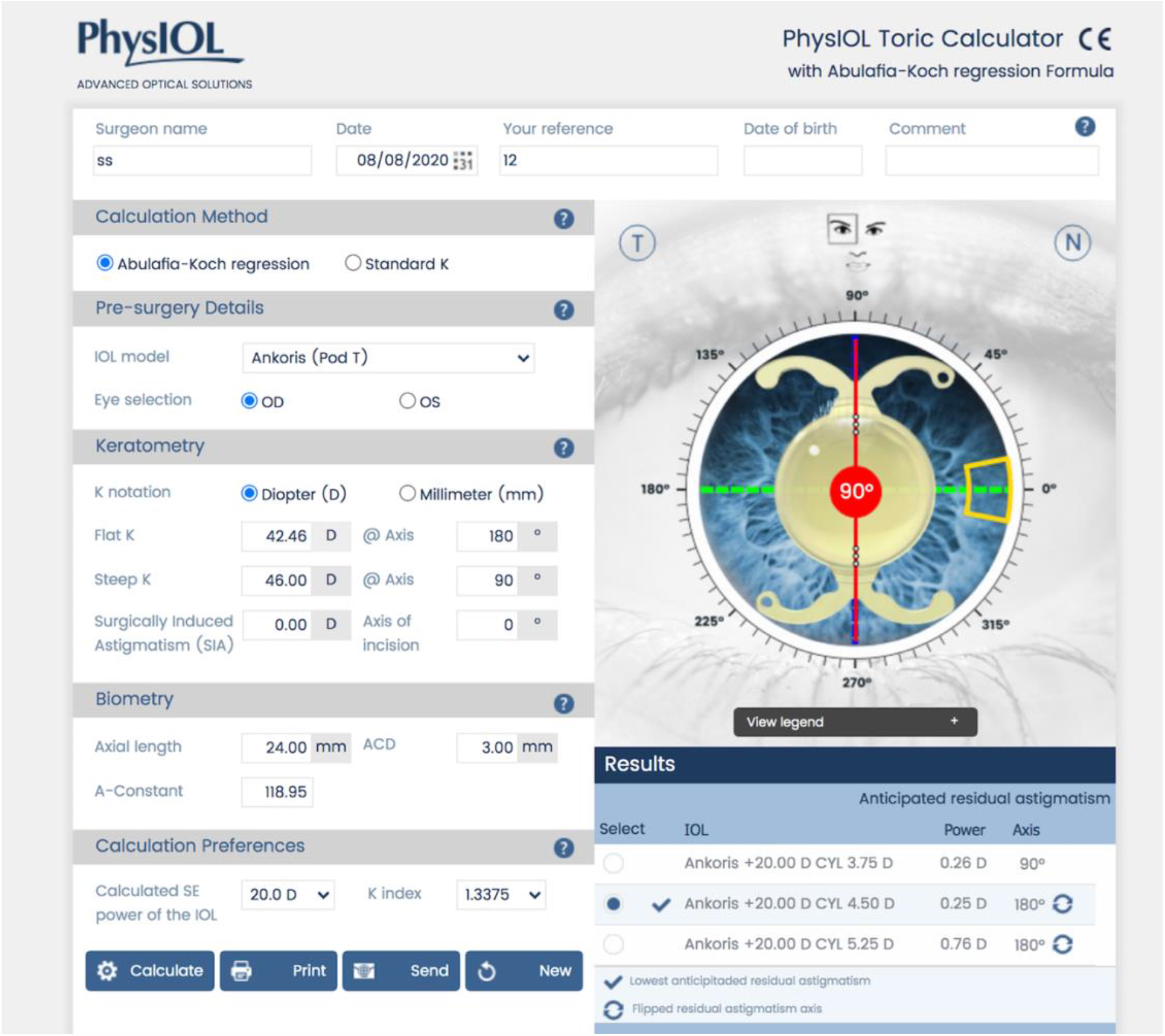

In this version of the calculator (on a slightly different example) and because of the inclusion of a contribution of posterior astigmatism (Abulafia-Koch regression), it is not immediately clear what is the amount of astigmatism being corrected. The methodology outlined can nevertheless be applied.

1. Here (*H*, Ω, *B, U*) = (4.50,0.25,3.*7*5,0.26) diopters
2. Determine ***α*** = Ω + *U* = 0.25 + 0.26 = 0.51, the *dioptric step at the corneal plane*
3. Calculate *n* by 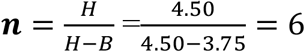
4. Estimate ***χ*** based on surgical experience or all contributions to misalignment, the calculate 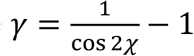 or use 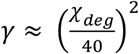. We take ***χ*** = **8**°. 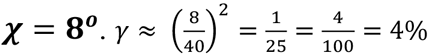
5. Substitute the values obtained in 1) to 3) in 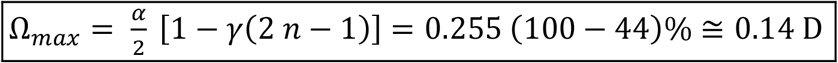
6. Overcorrect with *H* if and only if Ω < Ω_*max*_, otherwise choose *B*. Since Ω = 0.25 *D* and Ω_*max*_ = 0.14 *D* we clearly have Ω > Ω_*max*_ and overcorrection should be avoided and undercorrection selected.

We can also apply the *A*_*min*_ method with *n* = 6

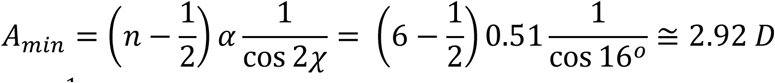

And compare to 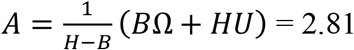 and because *A*_*min*_ > *A* the overcorrection is not recommended

We can verify *A*_*min*_+ Ω_*max*_ ≅ *h* = *nα* as we would expect. Indeed, we have *A*_*min*_ + Ω_*max*_ ≅ 2.92 + 0.14 = 3.06 = 6 × 0.51.

We can also determine the crossing angle for the values chosen by the calculator by applying *Eq* (*χ H*Ω*BU*). We find *χ* ≅ 1.*7*°. Once again this may be an unrealistic expectation in most surgical cases.

### ALCON split-the-difference with posterior corneal astigmatism inclusion

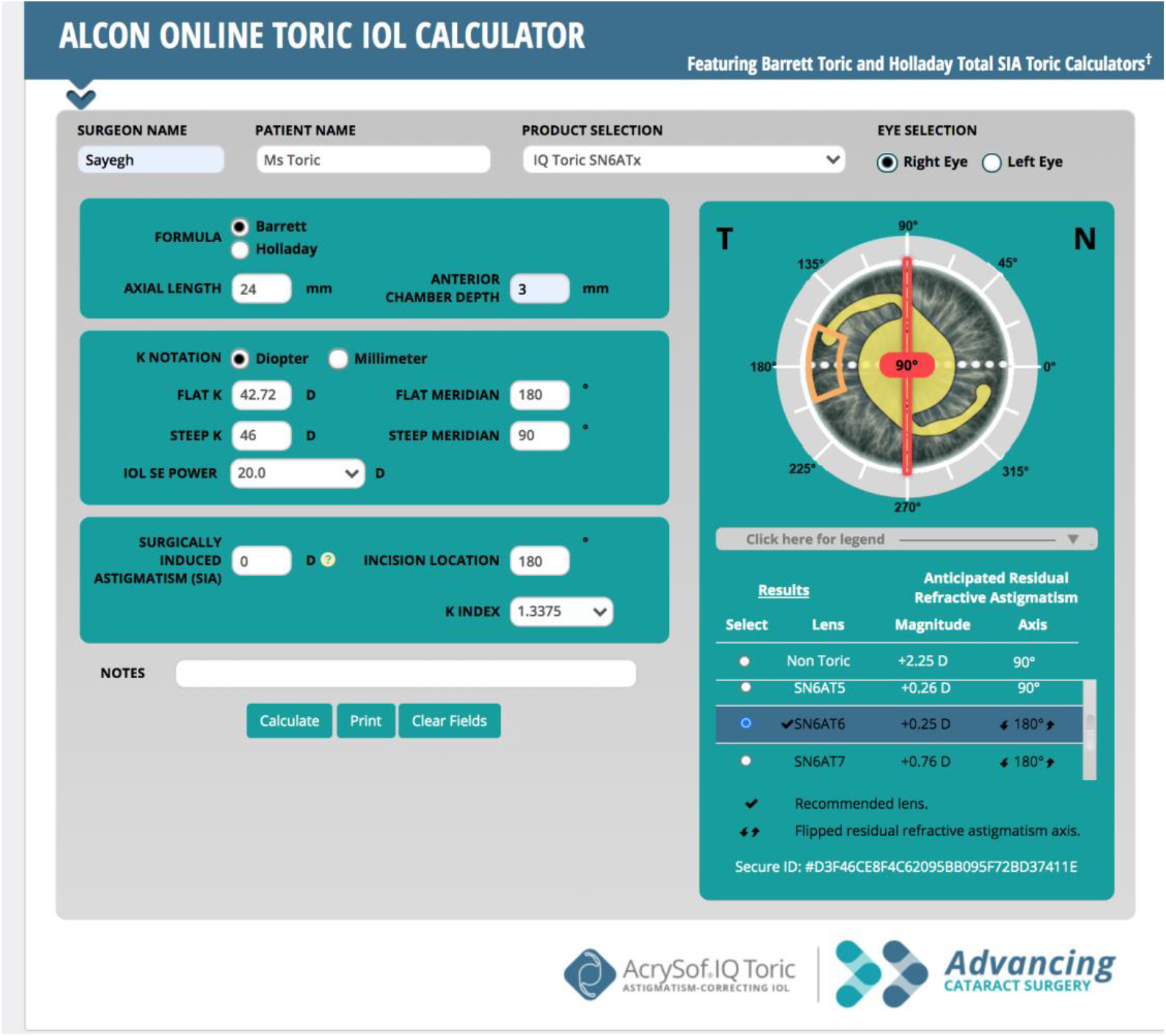

This example is identical to the one first used for PHYSIOL and the step, as well as the toricity ratio used by both manufacturers are the same. Our main interest is to show that there is an overcorrection of 0.25 D selected in both cases. In this case with a drastically different selection of a T6, the value of n is 5. Notice also that we are given a value of the astigmatism residual in case of a non-toric which presumably coincides with the computed total corneal astigmatism once all contributions including posterior astigmatism are included. With n = 5 we have O_max_[5, 0.75, 1.47, 8] = 0.16 D

Where O_max_ computes Ω_*max*_ as a function of the variables *n, σ, τ, χ* in that order. Once again, we can see that the suggested correction with the split-the difference approach as provided by the manufacturer, appears to overestimate the maximum overcorrection for realistic cases. Here too, while not reproducing other IOL calculators by the same manufacturer we point out the fact that the suggested tIOL for the same eye has a different value even though it also overcorrects by a larger amount than the one we suggest. Indeed the “Holladay Total SIA Toric” suggests a T7 tiOL with an overcorrected “flipped” residual of 0.18 D.

We can also determine the crossing angle for the values chosen by the calculator by applying *Eq* (*χ H*Ω*BU*). We find *χ* ≅ 1.9°. Again, this is unrealistic in most surgical cases.

### Johnson and Johnson split-the-difference with posterior corneal astigmatism inclusion

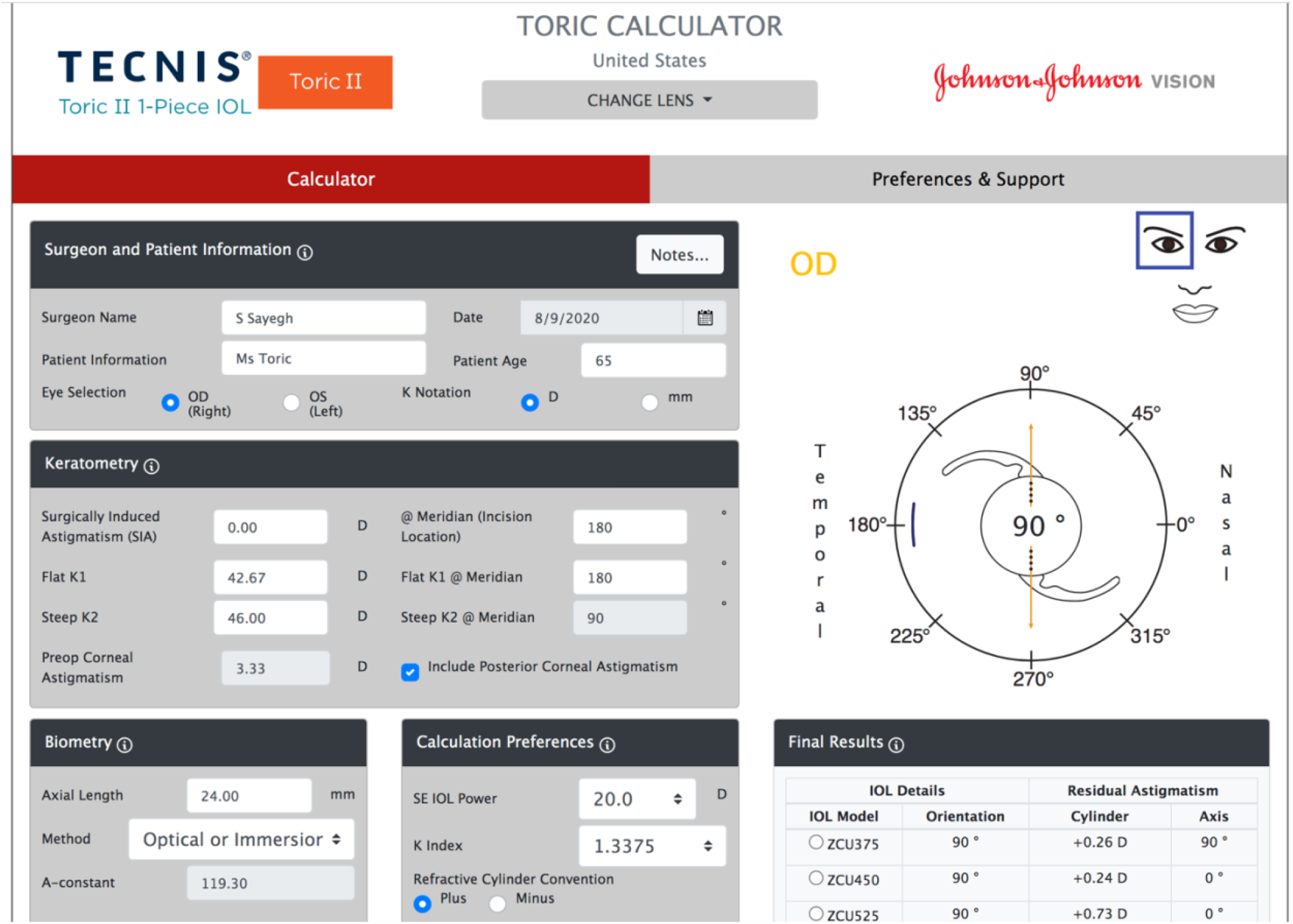

The values for the new ZCU tIOLs of JJ are essentially identical to those of Alcon Tn series (see Table 1). Calculation yields three possible solutions with the middle one presumably the recommended one. We can see in this example too (n = 6) that the split-the-difference approach is used yielding 0.24 D overcorrection while the recommended O_max_[6, 0.75, 1.50, 8] = 0.14 D. Appendix C illustrates how to deduce the total corneal astigmatism and toricity ratio from the output given. We can easily calculate A = 2.76 D. We know 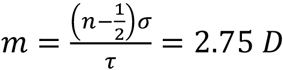 and 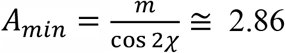*D* for an angle *χ* = 8°. Clearly *A*_*min*_ < *A*_*min*_and the overcorrection should be rejected. Similarly, we calculate

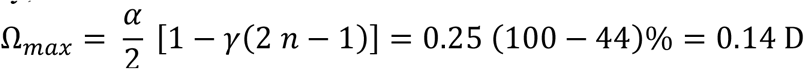

and verify *A*_*min*_+ Ω_*max*_= *h* = *nα* as we would expect. Indeed, we have *A*_*min*_+ Ω_*max*_≅ 2.86 + 0.14 = 3.00 = 6 × 0.50.

We can also compute the cross angle *χ* where the intersections of the overcorrection residual and undercorrection residual occur.

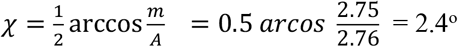 so in order to accept the overcorrection provided by that calculator we should be confident that misalignment from all causes will not exceed about 2 degrees, a very unlikely eventuality as previously discussed.

### ZEISS

ZEISS, while following the same general structure as exemplified by the above manufacturers, has a few significant differences that are worth noting before presenting an example of the methods they use to handle overcorrection.

The value of the step is *σ* = 0.50 *D* rather than the more common 0.75 D. Zeiss also provides a wider range of astigmatism correction in its tIOLs (up to 12 D of cylinder), corresponding to extending the values of n well beyond the value of 8 we have dealt with previously.

Initially, Zeiss did not overcorrect^1^ but fielded a newer calculator in mid 2020 which is the one we use here (ZCALC 2.2.0) It does now use overcorrection in the manner we will illustrate. This will also illustrate how to deal with a calculator that returns a single choice for the toric IOL, that choice itself being possibly associated with an overcorrection.

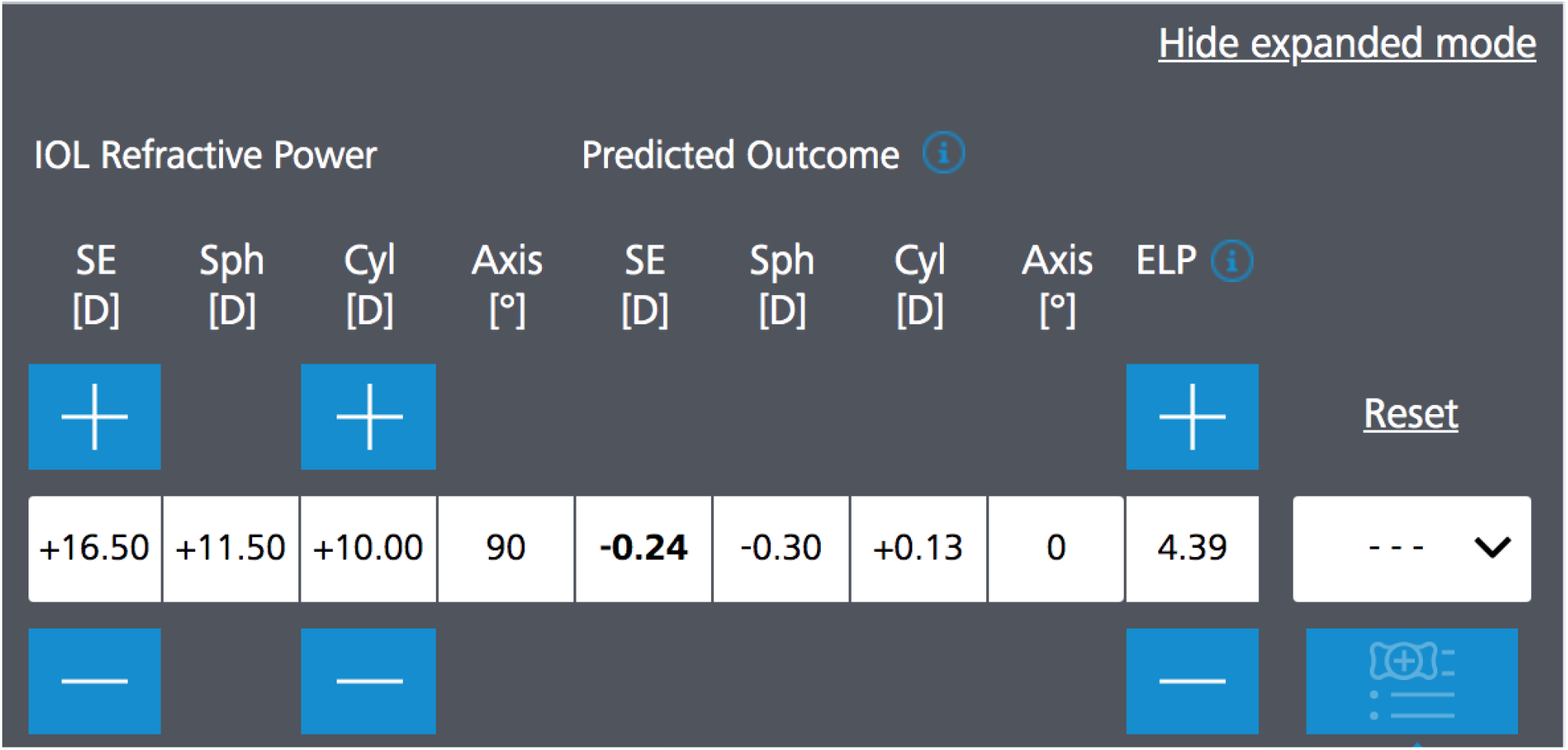

We will try to ask two questions in the context of what may be an extreme example that illustrates how the options for overcorrection narrow severely for high astigmatism. The above solution offered by ZCALC 2.2.0 is for values:

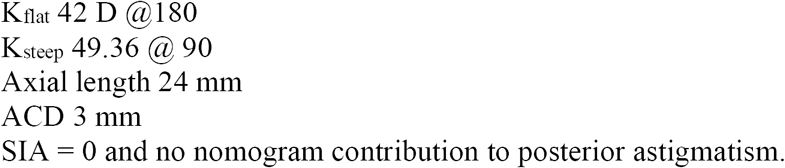

We are attempting, therefore, to correct 7.36 D of astigmatism. *A* = *7*.36 *D* We choose these values to focus the discussion on the main emphasis of overcorrection and whether it should or should not be allowed.

From the returned solution in the Zeiss screenshot above we can see that *H* = 10 *D*, n = 20, Ω = 0.13 *D*

Using the Sayegh-Gabra formula^5^ we determine *τ*:

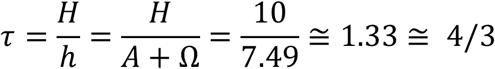

We now have all the parameters needed to compute

1. The min astigmatism that should be corrected for a given misalignment, say 8 degrees
2. The maximum overcorrection that should be allowed for a given misalignment, say 8 degrees
3. The angle *χ* at which overcorrection would become less advantageous than undercorrection if we were to accept an overcorrection for the given value of A

In sequence we have

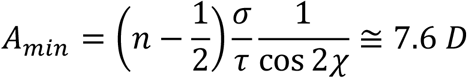

Notice that this value exceeds *h* = *7*.49 *D*! This means that for an angle *χ* = 8° of misalignment there is no degree of overcorrection that is favored, no matter how small.

Computing the maximum overcorrection, we expect a similar result. Indeed the computation of O_max_[20, 0.50, 1.334, 8] yields a negative result, again signaling that there is no allowable overcorrection for that degree of misalignment.

The simple physical interpretation of course is that the intersection of the overcorrection and the undercorrection curves has happened earlier than 8 degrees, just as represented on Figure 3.

To determine what is the angle of intersection we use

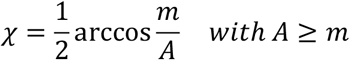

We find *χ* ≅ 3.6 < 4°

So before a misalignment of 4° is reached, the overcorrection “advantage” will be lost. Until we are able to operate with that level of confidence in the alignment of the tIOL with the correct value and axis of the total corneal astigmatism, no such overorrection should be attempted for that level of astigmatism.

## DISCUSSION AND CONCLUSION

In the first part of this trilogy we showed that in case of mismatch between an overcorrecting tIOL and the astigmatism it is intended to correct, the *angle of doom Δ*, is reached much sooner than the commonly taught 30 degrees. This is reflective of the fact that in the case of increasing misalignment with overcorrection, the residual astigmatism grows significantly faster than it does in the case of an undercorrection. Exact equations and appropriate approximations were given. Within that same paradigm we demonstrate here that there are profound implications regarding the optimal overcorrection that should be attempted when perfect alignment cannot be excluded, which occurs in the vast majority of realistic surgical cases. We give exact results and methods as well as practical surgical approximations that can be taken to the OR and applied to the next generation of toric IOL calculators. We have demonstrated the approach with examples, some which demonstrate suboptimal tIOL choices by current calculators. Part three of this trilogy I dedicated to a detailed discussion of a number of additional examples. This work and its associated methods promise to improve the surgical results in cataract refractive surgery with tIOLs for a majority of our patients.

## Data Availability

All necessary data exists within the manuscript.

## Appendix *A*

### γ has a simple clinical interpretation

From Equation (*γχ*) and writing λ for *χ* it is easily shown that

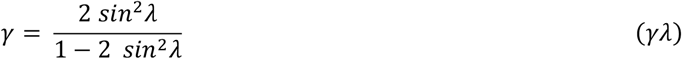

We thus have

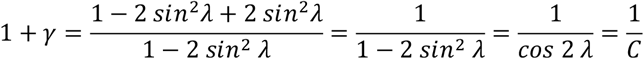

Equation A reads 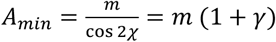

Therefore, we have

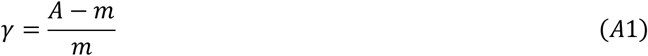

In other words, *γ is the fractional excess of A with respect to the midpoint* 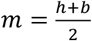

### γ has a simple clinically useful approximation

This equation relates λ and *γ* directly and provide clinically useful computational approximations: Note that for λ = 0, we will have *β* = *γ* = 0,*A* = *m*and Ω = *U*, as would be expected and shown in Fig[2]. Inverting Equation (*γ*λ) we obtain

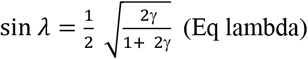

With

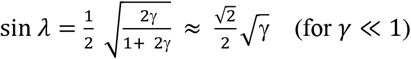

Using the small angle approximation *sin* λ ≈ λ and expressing the result in degrees, we obtain the very good approximation

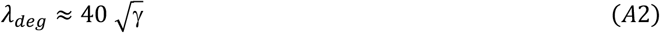

Or

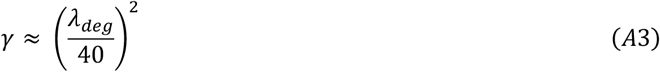

Note, as indicated in table [A1], that this approximation is excellent for the clinically relevant range of variables, as λ_*deg*_ will not extend beyond 15 degrees in most clinical scenarios, and the approximate form can thus be used for rapid mental evaluation of undercorrection or overcorrection choices.

| $\lambda_{deg}$ | $\gamma$ from exact expression $(\gamma\lambda)$ | $\gamma$ from approximation (A2) |
| --- | --- | --- |
| 0 | 0 | 0 |
| 5.0 | 0.0154 | 0.0156 |
| 10. | 0.064 | 0.063 |
| 15. | 0.15 | 0.14 |

Table [A1] Relationship between exact and approximate values of *γ,*the fractional excess astigmatism relative to the midpoint, as a function of *λ* _*deg*_,the angle at which overcorrection residual equalizes undercorrection residual. First differing decimal is displayed.

## Appendix *B*

Here we determine λ = *χ*, the angle at which the overcorrection residual crosses that of the undercorrection as a function of A. (*Eq χ*) reads

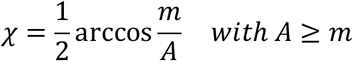

Writing 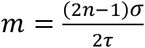 with 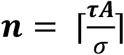 we can now plot *χ* as a function of A

For A from 1 D to 4 D for a given selection of *σ* and *τf*°*r* example with *σ* = 0.*7*5 and *τ*= 1.5, values most commonly encountered in clinical practice.

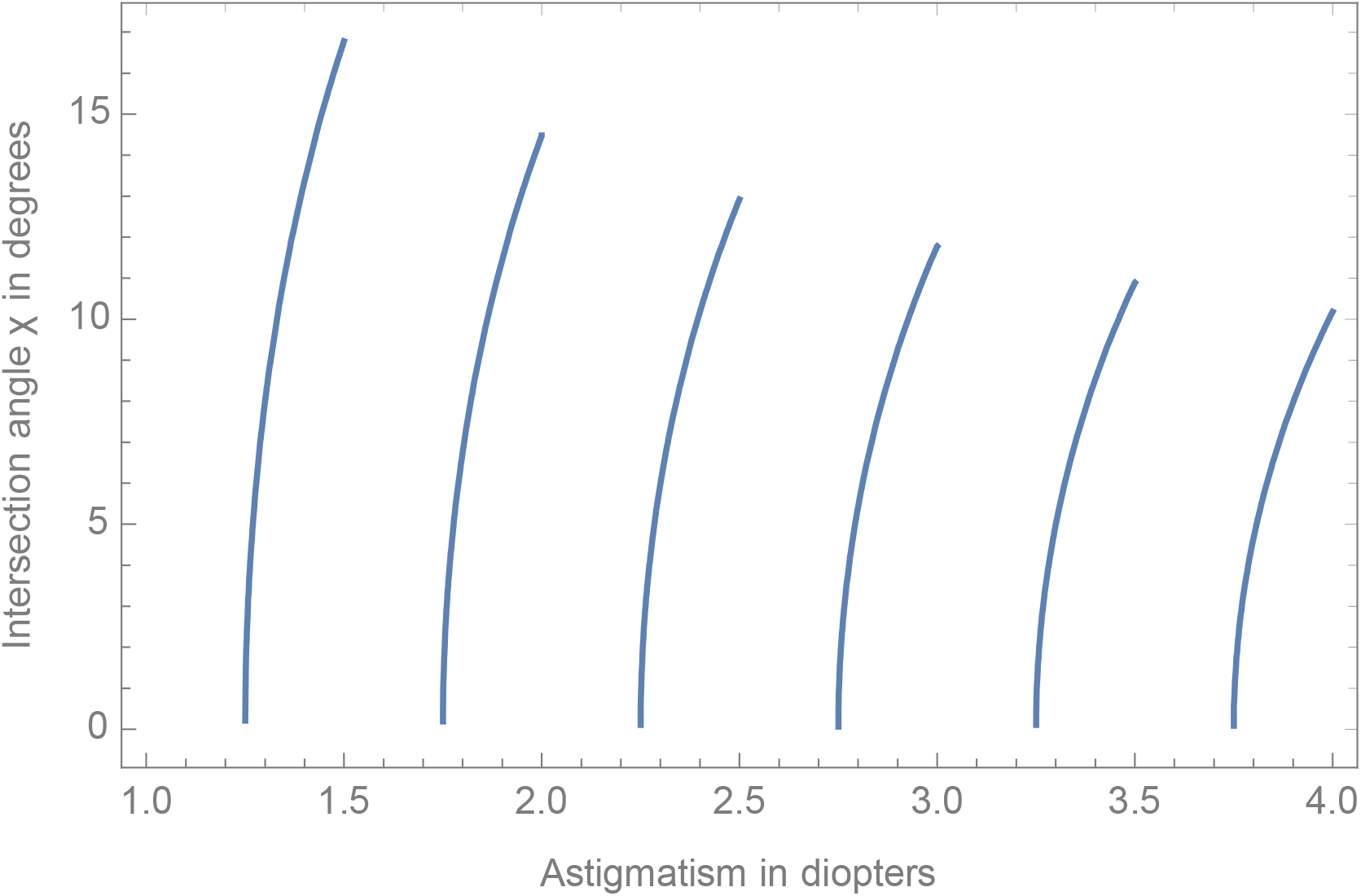

*Figure B1 For Tn sequence σ* =*0*.*7*5 *and τ* = 1.5, *we plot the angle of intersection χas a function of astigmatism to be corrected*

Note that when undercorrection < overcorrection we get no real solution for the angle, as we should expect! Also note that *χ* can be approximated by 40 √*β* with 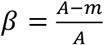

So, if A is very close to mid-point m, *χ* will be very small (this should be clear since when A = m, overcorrection and undercorrection are equal and “intersection” is at zero as the curves arise from the same point at zero misalignment)

So very small differences favoring overcorrection are likely to disappear under minimal misalignment! The effect depends on m/A …so there is a step like behavior modulated by A.

## Appendix C

Reading the T leaves: How to deduce toricity ratio and total corneal astigmatism even if you are not told.

Sayegh showed how to calculate the toricity ratio and examples were given as Sayegh-Gabra formula. We extend these results in the following to allow to also obtain the total corneal astigmatism in many instances where it is not given explicitly,

From Figure 2 we can see

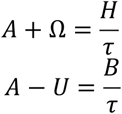

These two equations can be written in matrix form

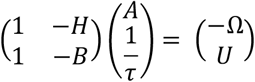

Both A and *τ*can be determined by inverting the matrix. This is done by inspection by exchanging the diagonal elements and flipping the signs of the non-diagonal elements and dividing by the determinant *H* − *B* = *σ*. The results are as follows

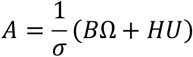

And

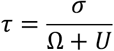

We demonstrate on the JJ example where we know *σ* = 0.*7*5 and we had

|  | Cylinder of tIOL in D | Residual in D |
| --- | --- | --- |
| Undercorrection | B= 3.75 | U = 0.26 |
| Overcorrection | H = 4.50 | $\Omega = 0.24$ |

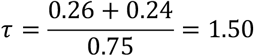

A is obtained by cross multiplying as per equation above We get 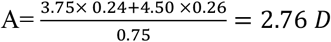

Note that when given more than two options for correction along with the corresponding residuals it is possible to determine *A and τ*either by selecting a pair of equations as above or, if the equations are not linearly dependent, by solving the overdetermined system of equations, for example with a Moore Penrose pseudo inverse.

## Guide to symbols and key equations

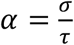 “step” increment in diopter between two successive tIOLs, at the corneal plane

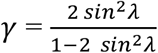 is also is the fractional excess of A with respect to the midpoint *m*(See Appendix)

Δ “angle of doom” at which astigmatism correction is nullified

λ angle of misalignment *from all causes* between the tIOL and the astigmatism to be corrected

*σ* “step” increment in diopter between two successive tIOLs, at the IOL plane

*τ*toricity ratio, conversion factor from cylinder at IOL plane to cylinder at corneal plane

*χ* crossing angle of overcorrection and undecorrection residuals as a function of λ

*ω* overcorrection relative to *A*

Ω overcorrection in diopters, at the corneal plane

A astigmatism to be corrected, at the corneal plane

b cylinder of undercorrecting tIOL at the corneal plane

*B* cylinder of undercorrecting tIOL at the IOL plane

*C* = cos 2λ

*h* cylinder of overcorrecting tIOL at the corneal plane

*H* cylinder of overcorrecting tIOL at the IOL plane

*L* cylinder of generic tIOL at cornea plane (See Figure 1)

*m* midpoint between *h* and *b*, mean or midpoint of *h* and *b*

*n (possibly* integer) value determining pair of under and overcorrecting tIOL,*if integer* 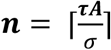

*R* Generic residual astigmatism (See Figure 1)

*u* undercorrection relative to A

U Undercorrection in diopters, at the corneal plane

**“First Quantization”**

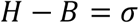

**“Second Quantization”**

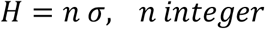

**Overcorrection limit**

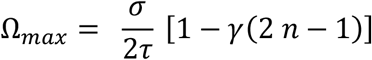

## Example of a Method

1. Determine *σ* by selection of manufacturer’s sequence of tIOLs.
2. Determine *τ* by measuring the eye’s axial length and K values using an appropriate formula or nomogram^45^
3. For astigmatism *A*, calculate 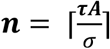
4. Select corresponding undercorrecting (*n* − 1)*σ* and overcorrecting *nσ* candidate tIOLs.
5. Estimate *χ* based on surgical experience or on all contributions to misalignment, then calculate 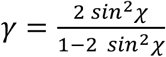 or the approximation 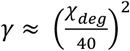
6. Substitute the values obtained in 1) to 5) in 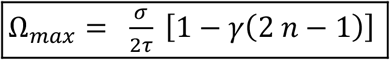
7. Overcorrect A with *H* if and only if 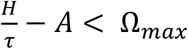,otherwise choose *B* = *H* − *σ*

## Footnotes

1 ^1^Note that if we use 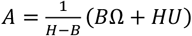 with the values given we obtain A = 3.32 D instead of 3.28 D. This is due to rounding used in the calculator under examination. Indeed a function Atot[H_, O_, B_, U_] := (H U + B O)/(H - B) returns 3.32 for Atot[5.25, 0.25, 4.50, 0.26]. However Atot[5.25, 0.248, 4.50, 0.256] returns 3.28 as should be while the entries 0.248 rounds to 0.25 and 0.256 rounds to 0.26. With a value of *A_min_* = 3.45 this discrepancy clearly does not distract from the main conclusion but may need to be taken into consideration in borderline cases. It is fully analyzed in Part 3 which is dedicated to implementation and numerical examples.

